# Innate immunity associates with protection from pneumococcal colonisation, but colonisation does not confer capsule-independent protection

**DOI:** 10.64898/2026.06.23.26355871

**Authors:** V. Connor, E. Mitsi, K. S. Cheliotis, E.L. German, P. Gonzalez-Dias, S. Pojar, E. Nikolaou, S.P. Jochems, S.H. Pennington, C. Hales, A. Hyder-Wright, H. Adler, S. Zaidi, J. Reiné, S.B. Gordon, H. Hill, E.N. Miyaji, A. M. Collins, R. Malley, Y.J. Lu, R.O. Tostes, C.P. Jewell, C.M. Weight, B. Urban, C. Solórzano, J. Rylance, D.M. Ferreira

## Abstract

Nasopharyngeal colonisation with *Streptococcus pneumoniae* is a prerequisite for transmission and disease and represents an important immunising event. While colonisation induces serotype-specific immunity, the mechanisms underlying heterologous protection remain unclear. We developed a controlled human infection model using pneumococcal serotype 15B and investigated colonisation dynamics, immunogenicity, and cross-protection against subsequent heterologous challenge with serotype 6B.

Fifty-four healthy adults were intranasally inoculated with 15B at escalating doses. Colonisation rates peaked at 31.4% with 8 × 10⁴ CFU per naris, lower than those historically observed with 6B and 3 strains. Density was also lower than previously observed with other strains. *In vitro* assays demonstrated that 15B adhered more readily to epithelial cells than 6B, but was less efficiently internalised, potentially reducing attack rates and colonisation density. Colonisation with 15B induced capsular polysaccharide-specific serum IgG, but baseline humoral immune measures did not predict protection from acquisition. Prior colonisation with 15B did not reduce acquisition of 6B upon re-challenge.

Analysis of nasal microbiopsy samples revealed distinct innate activation signatures. Resistance to colonisation was associated with elevated baseline MIP-1α and MIP-1β responses upon *in vitro* stimulation, whereas carriage was associated with enhanced chemokine and IL-6 responses. Local innate immune activation, rather than circulating antibody responses alone, may therefore contribute to colonisation control.

We demonstrate that experimental colonisation with 15B does not confer heterologous protection against 6B and highlight the importance of mucosal innate immune conditioning in serotype-independent defence. Strategies enhancing nasal innate immune recruitment and activation may be required for broader protection against pneumococcal colonisation.

## Introduction

*Streptococcus pneumoniae* (pneumococcus) is a bacterial pathogen that commonly colonises the upper respiratory tract. Nasopharyngeal colonisation is common with reported point prevalence of 40-90% in children and 10-25% in adults (1–3). Nasopharyngeal colonisation is the primary reservoir for pneumococcal transmission in the community and is a pre-requisite for progression to invasive pneumococcal disease (4, 5). Pneumococcus is a leading cause of bacterial pneumonia, meningitis and otitis media, which are associated with significant morbidity and mortality worldwide (6, 7); in 2024, the World Health Organisation (WHO) named *S. pneumoniae* as a priority pathogen once again due to the level of threat to human health (4, 8).

Rates of pneumococcal acquisition and colonisation vary greatly by age, geographical location and socioeconomic settings (9). Pneumococcal colonisation has also been found to be an important immunising event in healthy adults (10, 11). During childhood, repeated exposure to pneumococcus supports the development of immunity that associates with declining rates of colonisation and disease into adult life (12, 13). Immunity acquired through natural pneumococcal colonisation is likely to be only partially serotype-specific and is complemented by broader, non-capsular immune responses that act across serotypes (13). Suboptimal serotype-specific immunity allows repeated colonisation events and supports the coexistence of multiple pneumococcal serotypes in human populations (13). In murine models, antigen-specific B-cell and T-cell responses develop during colonisation and protect against subsequent re-colonisation (14–17).

In experimental human models, colonisation has been shown to increase serum IgG and secretory IgA levels against pneumococcal proteins as well as capsular polysaccharide of the challenge strain in the periphery and in the nasal mucosa (11, 18, 19). Experimental colonisation also induces pneumococcal-specific IL-17A producing CD4^+^ T cells in both blood and lung mucosa (12).

Earlier work demonstrated that experimental human colonisation with serotype 6B protects against re-acquisition of the same pneumococcal strain for up to 11 months after the first colonisation episode in 70-100% of study participants (11). Protection against re-acquisition was associated with increased levels of IgG to several pneumococcal proteins and capsular polysaccharide mounted in response to the initial carriage event (11).

Here, we report the development of a controlled human infection model (CHIM) for pneumococcal serotype 15B in healthy adults and its use to determine whether immune responses can provide protection to subsequent heterologous colonisation. 15B was not included in the 13-valent polysaccharide conjugate vaccine used in the UK but is an important serotype in terms of disease burden and its association with antibiotic resistance. 15B has hence been included in the 20-valent vaccine (20, 21).

We used this novel 15B CHIM combined with our established 6B CHIM to understand cross-protection generated by colonisation-induced immunity. After examining B-cell mediated immunity and IgG levels against pneumococcal proteins and capsular polysaccharide, we observed that the serotype-independent immunity induced by 15B colonisation was suboptimal and therefore, did not associate with protection against heterologous-serotype rechallenge. On the other hand, the status of nasal mucosa activation and its ability to signal monocytes and neutrophils (MIPs, IL6) suggest that innate immune responses may play an important role in protection against colonisation.

## Methods

### Statement on sex as a biological variable

We did not disaggregate data by sex due to limited sample sizes.

### Recruitment

We recruited healthy adults aged between 18 and 50 years of age. All participants were non-smoking adults with no history of systemic or respiratory disease. As per our standard human challenge study protocols (11), we excluded individuals who had close regular contact with those at risk of pneumococcal infections such as children under 5 years of age and immunocompromised adults.

### Study approval

All participants gave informed written consent. Ethical approval was obtained from the National Health Service Research Ethics Committee, Liverpool East (15/NW/0931) and the trial was registered with ISRCTN (Trial Registration Number: ISRCTN68323432).

### Study protocol and sample collection

Pneumococcal stock preparation and inoculation were performed as previously described [19]. Briefly, volunteers were inoculated with serotype 15B *S. pneumoniae* intranasally (clinical isolate 15B P1262. European Nucleotide Archive accession number: ERS2632437). Participants were challenged with 2 × 10^4^, 8 × 10^4^ or 16 × 10^4^ colony forming units (CFU) per naris in 100 μL saline (Supplementary Figure 1). For re-challenge experiments, those participants who became experimentally colonised with 15B were invited to undergo re-challenge with serotype 6B (Clinical isolate 6B BHN418. GenBank accession number ASHP00000000.1). Re-challenge was performed 5-9 months following initial challenge (using serotype 6B at 8 × 10^4^ CFU per naris in 100 μL saline).

Pneumococcal colonisation was determined using nasal wash, as previously described (22). Nasal wash, peripheral blood and nasal microbiopsy (ALS Rhino-Pro^©^, Arlington Scientific) samples were acquired according to a standardised sampling schedule (Figure 1).

**Figure 1.**
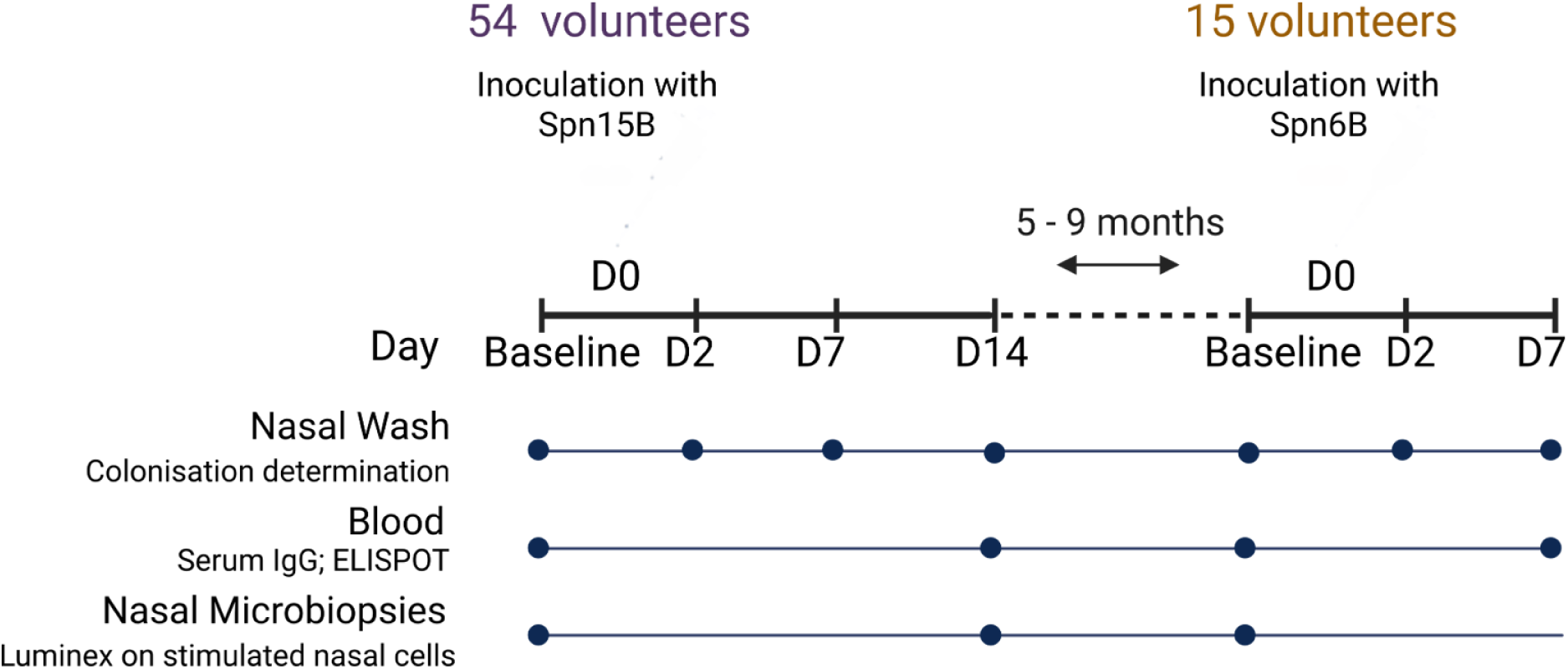
Study sampling schedule. Baseline nasal wash samples were obtained from all participants up to 1 week before inoculation and on days 2, 7 and 14 post inoculation for initial challenge. For re-challenge experiments, samples were collected up to 1 week prior to inoculation and on days 2 and 7 post second inoculation. Blood samples were obtained from participants up to 1 week before inoculations and at the final follow up visits (day 14 for initial challenge and day 7 for re-challenge). Nasal microbiopsies (ALS Rhino-Pro©, Arlington Scientific) samples were also obtained at baseline for challenge and rechallenge and at 14 days post initial challenge.

### Determination of colonisation

Pneumococcal colonisation and density were assessed in all nasal washes by microbiological culture, as previously described (22). The serotype recovered was determined using a latex agglutination kit (Statens Serum Institute, Denmark). Participants were classed as carriage-positive if the inoculated serotype of pneumococcus (15B for initial challenge and 6B for re-challenge) was detected in any nasal wash taken after inoculation by culture.

Nasal washes were also assessed for pneumococcal colonisation and density using *lytA* quantitative polymerase chain reaction (qPCR). Bacterial DNA extraction and quantification of pneumococcal DNA by qPCR were performed as previously described (23).

### Anti-Pneumococcal Capsular Polysaccharide IgG determination

Anti-capsular polysaccharide (PS) IgG levels to serotype 15B were measured by the standard World Health Organisation ELISA method, according to established protocols (24). Briefly, serum samples were incubated with cell whole polysaccharide (CWPS) then were added to plates pre-coated with type-specific capsular polysaccharide. Antibody levels were detected using a goat anti-human IgG conjugate and p-nitrophenyl phosphate. Optical density was measured at 405nm and 690nm. Reference serum 89-SF was used for standard curves. All samples were analysed in duplicate.

### Anti-protein IgG determination by Luminex

As described elsewhere (25), seventy-five pneumococcal protein antigens were conjugated to magnetic microspheres at a concentration of 5 µg per 1 × 10^6^ microspheres. Microspheres were diluted 1:5 in 1% BSA–PBS to achieve 50 beads/µL and 25 µL were added to each well of a 96-well plate, followed by 25 µL of serum diluted 1:20,000 in assay buffer. Pooled serum diluted 1:50 in assay buffer served as a positive control, and assay buffer as a negative control. Plates were incubated overnight at 4 °C in the dark with shaking (300 rpm). After three washes, 50 µL biotinylated anti-human IgG Fc (2 µg/mL) was added and plates were incubated for 30 min at room temperature with shaking, followed by three washes. Streptavidin-PE (2 µg/mL, 50 µL) was then added and plates incubated for 10 min. After three further washes, 50 µL assay buffer was added and plates were read on a Luminex LX200. All samples were analysed in duplicate.

### Enzyme-linked Immunospot Assay for Memory B-Cells

Enzyme-linked immunospot assay (ELISPOT) plates were prepared, as previously described, with the following modifications: isolated PBMCs were used from frozen, thawed and seeded at 5 × 10^5^ per well and stimulated with R848 and IL2 for 5 days. Cell harvesting and seeding on the ELISPOTs plates coated with 15B purified capsular was performed, as previous described (26).

### Luminex analysis of stimulated nasal cells

Stimulation of nasal cells from nasal microbiopsies (ALS Rhino-Pro^©^, Arlington Scientific) was carried out with the heat-killed pneumococcus 15B or 6B or RPMI cell culture medium (mock stimulation) for 16 hours as previously described (27). Levels of IL6, IL10, GM CSF, TNFα, MIP-1β and MIP-1α were measured in cell supernatant by a Human Custom Procartaplex 6-plex magnetic Luminex cytokine kit (ThermoFisher). Data were acquired on a LX200 and analysis was carried out using xPonent 3.1 software, according to the manufacturer’s instructions. All samples were analysed in duplicate.

### Adhesion/invasion assay with Detroit 562 cells

Detroit 562 epithelial cells were seeded in 12-well plates and cultured to confluency, usually 7-9 days post plating. Cells were maintained in alpha MEM supplemented with 10% Foetal Bovine Serum (FBS, Gibco). Medium was replaced with alpha MEM containing 1% FBS for infection assays. Cells routinely tested negative for mycoplasma. *S. pneumoniae* stocks frozen at optical density 0.3 were thawed, centrifuged at 8000 × g for 8 min, and resuspended in 1% FBS in alpha MEM. Bacteria were added to triplicate wells for each treatment condition. After 3 hours incubation, wells were washed three times with 1 ml Hank’s balanced salt solution (HBSS) to remove non-adherent bacteria. For internalisation assays, gentamicin (200 µg/ml in 1% FCS/MEM) was added after the initial wash and incubated for 1 hour to kill extracellular bacteria, followed by three further washes with HBSS. Cells were then lysed with 1% saponin for 10 min. Serial dilutions of lysates were plated in duplicate on horse blood agar to quantify colony-forming units (CFU) after 16 hours. Pre-inoculums and post-inoculums were recorded to monitor bacterial growth for the duration of the assay. Each experiment was repeated five times; four biological replicates were included in the final analysis following a change in inoculum preparation after the first experiment.

### Statistical analysis

For B cell data, statistical significance was determined via Mann-Whitney or Wilcoxon testing. Where appropriate, data were logarithmically transformed prior to statistical analysis. For adhesion/invasion assay data, statistical analysis was performed using two-tailed unpaired t-tests for inoculum and proportional data, and Mann–Whitney tests for adhesion and invasion assays.

Stimulated nasal cell data was cleaned prior to analysis as follows. Analyte measurements with a coefficient of variation (CV) between duplicates of >50% were excluded from analysis unless cytokine concentration was <10 pg/mL or greater than the value of the highest standard. Based on these criteria, a total of 45 analytes (8.5% of total acquired) were removed. Statistical analysis was carried out in RStudio (version 1.0.153). Statistical significance was determined via a Mann-Whitney or Wilcoxon test for unpaired and paired groups, respectively.

Raw anti-protein IgG detection data was cleaned prior to analysis as follows. The CV of replicates was calculated for all analytes. If any analyte had greater than 25% CV and both replicates had greater than 50 microspheres per well, the analyte was removed from analysis. If any paired samples had less than 50 microspheres of a given analyte in each well, the analyte was removed from analysis. Differences in IgG levels against pneumococcal proteins at baseline, were analysed using a ridge regression model fit using the “glmnet” package in RStudio (version 2024.12.0+467). To optimise model performance, 10-fold cross-validation was performed to determine the optimal lambda value that minimised the test mean squared error (MSE). The best lambda value obtained was 7.17. Model fit was assessed using McFadden’s R², calculated as 0.97, indicating a strong predictive performance. Receiver Operating Characteristic (ROC) analysis was performed to evaluate classification performance. To validate robustness, a 10-fold cross-validation procedure was implemented. AUC values were computed for each fold, yielding an average cross-validated AUC of 0.84, confirming strong model generalisability.

SP2070 and SP0084 were removed from longitudinal analysis of changes in anti-protein IgG due to >90% missing values. Longitudinal analysis was performed using RStudio (version 2024.12.0+467).

All laboratory analyses were performed blinded to colonisation status.

### Data availability

Data are available upon reasonable request by email directed to the corresponding authors at.

## Results

### Colonisation rates and densities over time

For the dose-ranging part of the study, 29 eligible participants were inoculated with 2 × 10^4^ (n=10), 8 × 10^4^ (n=10) or 1.6 × 10^5^ (n=9) CFU of 15B per naris. Whilst there was some acquisition of colonisation at each dose, colonisation rates were highest in the groups challenged with 2 × 10^4^ and 8 × 10^4^ CFU per naris (30% and 40%, respectively). Colonisation rates were lower at the highest dose (11% at 1.6 × 10^5^ CFU/naris). Based on these results, 8 × 10^4^ CFU per naris was used for the reproducibility part of the study, in which an additional 25 participants were inoculated. Total colonisation rate of all participants inoculated with 8 × 10^4^ CFU per naris (as determined by microbiological culture) was 31% (11/35) (Figure 2A). There were no reported significant adverse events related to the study (28). Cohort details for each inoculation dose are given in Supplementary Table 1.

**Figure 2.**
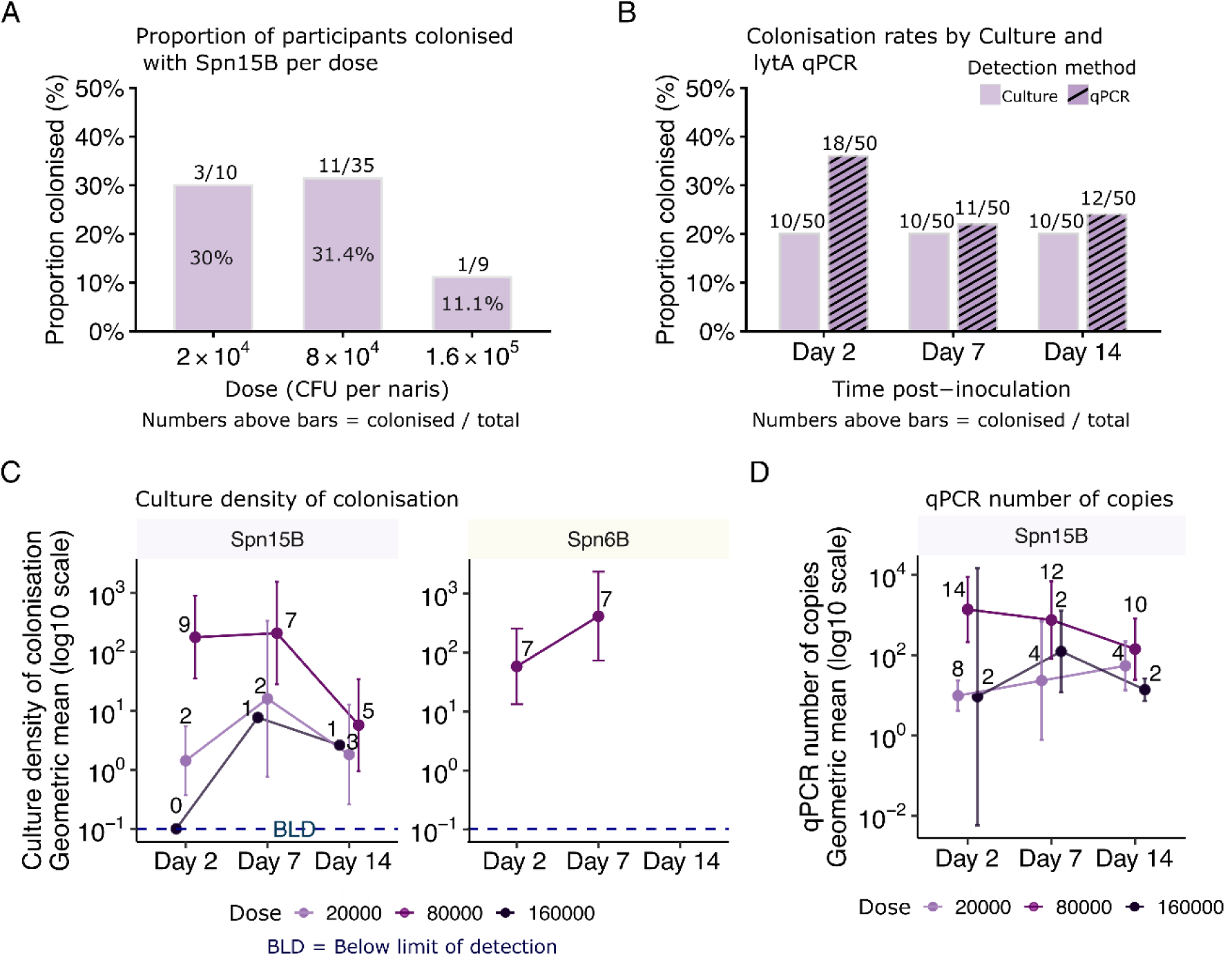
Colonisation rates and densities following experimental human pneumococcal challenge. (A) Colonisation rates with 15B per dose. For dose 8×10^4^ CFU/naris, dose-ranging and reproducibility cohorts were combined. (B) Colonisation rates with 15B determined by microbiological culture (violet) and by lytA qPCR (pattern). (C) Colonisation density with 15B (left, violet) and 6B (right, yellow) measured by microbiology culture. Points represent geometric mean density, and error bars indicate 95% confidence intervals. Lines correspond to inoculation dose: 2 × 10⁴ (light purple/lavender), 8 × 10⁴ (dark magenta/purple), and 1.6 × 10⁵ (very dark purple/near-black). The dashed blue line indicates the lower limit of detection. (D) Colonisation density with 15B measured by lytA qPCR. Bacterial density quantified by lytA qPCR is shown as geometric mean genome copies/mL with 95% confidence intervals at each time point. Participants determined to be naturally colonised with pneumococcus at baseline or during the study were excluded for this analysis. Differences across timepoints within each dose group were assessed using the Kruskal–Wallis test. No statistically significant differences were observed.

We excluded 4/54 (7%) of volunteers prior to analysis since they were identified as naturally carrying pneumococcus at baseline or at any point during the study. All 4 participants were carrying a serotype that could not be determined using the latex agglutination kit.

Two hundred nasal wash samples from the remaining 50 participants were tested for the presence of *S. pneumoniae* by *lytA* qPCR (in addition to culture) at the end of the study. Eight volunteers who had not been identified as being colonised by microbiological culture were identified as being colonised by qPCR (Figure 2B). Rates of experimental colonisation with 15B were similar at each time point by classical microbiology (Figure 2B), whilst rates determined by qPCR were highest at day 2 post inoculation (35%), then fell to 22% at day 7 and 24% at day 14 (Figure 2B). Colonisation density declined with time for volunteers inoculated with 8 × 10^4^ CFU per naris (Supplementary Figure 2). This drop was statistically significant between day 2 and day 14 when culture densities was assessed but not statistically significant when using qPCR density (culture; mean 20068 vs 116 CFU/ml nasal wash P= 0.03, Mann Whitney test) (Figures 2C, 2D). No significant change in densities was observed over time in either of the other two inoculum dose groups (Figures 2C, 2D).

### 6B is significantly more readily internalised by Detroit 562 cells compared to 15B

Given rates of colonisation and colonisation density were lower for 15B strain compared to 6B strain in the human challenge model, we evaluated differences in adherence and internalisation between these strains using well-established nasopharyngeal cell line (Detroit 562) in vitro assays (29).

Serotype 15B was significantly more likely to adhere (associate) to Detroit 562 cells than 6B after three hours of infection (Figure 3A). However, 6B was significantly more readily internalised by Detroit 562 cells compared to 15B (Figure 3B and 3C). Since we have previously shown that epithelium microinvasion occurs during colonisation (29), it is possible the lower cell internalization of 15B could contribute to the lower colonisation rates and density observed by this strain when compared with previous studies using 6B strain.

**Figure 3.**
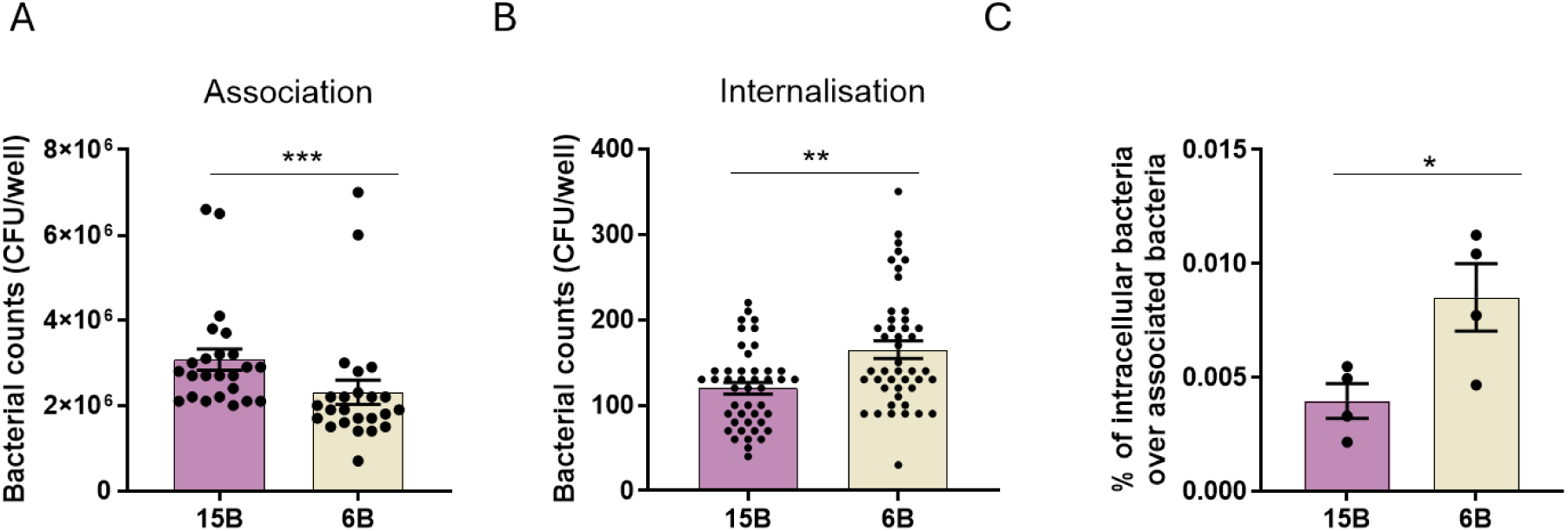
Number of 6B and 15B pneumococci. (A) associated with or (B) internalised by Detroit 562 cells after 3 hours’ incubation. (C) Proportion of intracellular over associated bacteria for 6B (yellow) and 15B (purple). **** indicates significant difference between 6B and 15B, P<0.0001, Mann Whitney Test. Bars indicate mean +/- SEM.

### Baseline 15B capsular polysaccharide (CPS) IgG levels do not associate with colonisation acquisition or control of density

We have previously reported that baseline levels of 6B and 3 CPS IgG do not correlate with protection against experimental colonisation by a 6B strain in unvaccinated participants (11, 26, 30). To determine whether this was also the case for 15B CPS, we measured IgG levels in serum samples from 34/35 participants (one was excluded due to natural colonisation at baseline) before inoculation with 8 × 10^4^ CFU per naris. Levels of 15B CPS IgG at baseline were similar between carriage-positive and carriage-negative participants (Figure 4A). Median baseline 15B CPS IgG level in carriage-positive volunteers was 3787 ng/mL (IQR: 1607-6465) compared with 3681 ng/mL (IQR: 1927-6008) in carriage-negative participants (*p*=0.91). Baseline levels of 15B CPS IgG were also not associated with the area under the curve of colonisation density over time assessed by either culture or *lytA* qPCR (Supplementary Figure 2).

**Figure 4.**
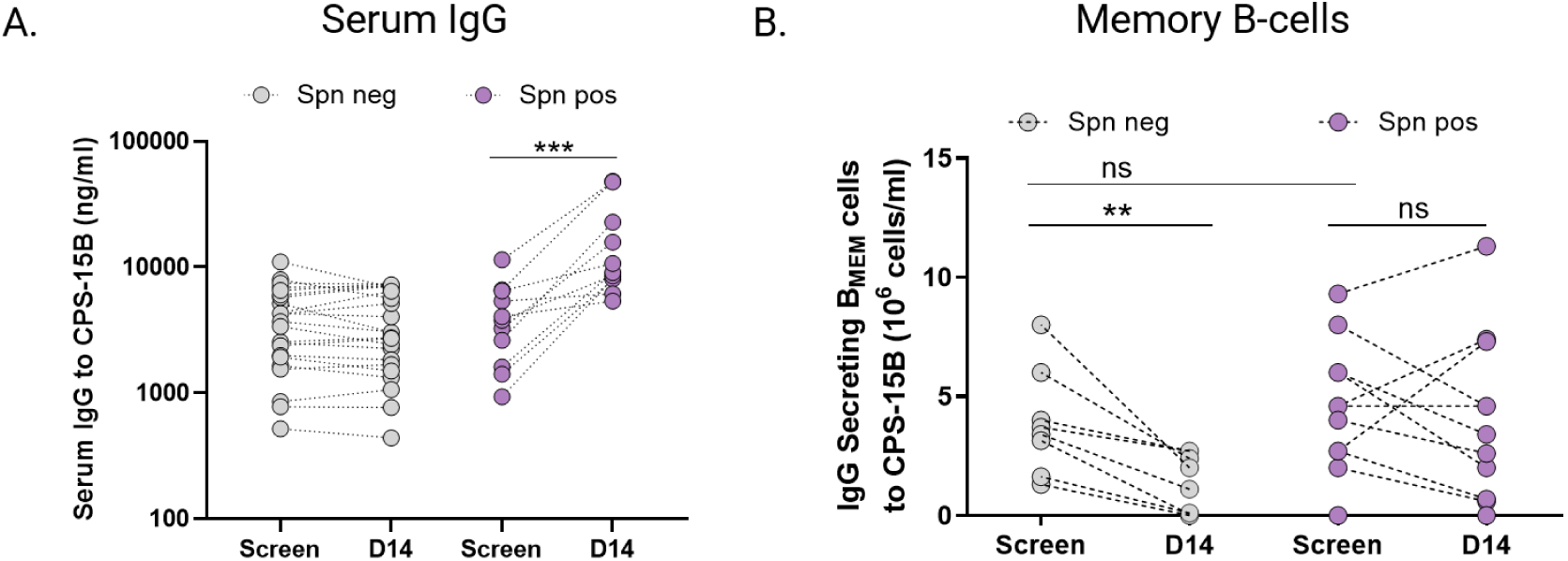
Humoral and memory B-cell responses to pneumococcal serotype 15B challenge. (A) Levels of serum IgG to 15B capsular polysaccharide measured at baseline and 14 days after pneumococcal challenge, determined by ELISA (all samples in duplicate) from carriage-negative participants (grey, n=23) and carriage-positive participants (purple, n=12). ***p<0.001 by Wilcoxon test. (B) Frequency of 15B-CPS IgG secreting B_MEM_ cells in carriage-negative (grey, n=7) and carriage-positive participants (purple, n=12) at baseline and 14 days post challenge with 15B. Enzyme-linked immunospot assays were performed against pneumococcal CPS 15B in blood samples collected from study participants before (−5) and after (day14) intranasal inoculation with live 15B. Wilcoxon test was used for comparisons within the same group, and Mann Whitney test for two groups comparisons. n.s: non-significant.

We have also shown that experimental colonisation with pneumococcus correlated with increased levels of CPS specific IgG levels in sera and this was dependent on the pneumococcal serotype and strain (19, 26, 31). To evaluate if experimental colonisation with 15B was immunogenic, we measured CPS specific IgG levels in paired baseline and day 14 serum samples. Carriage-positive participants had a 2.4-fold increase in levels of 15B CPS IgG at day 14 post inoculation compared to baseline (median 3787 ng/ml at baseline vs 9067 ng/ml at day 14, *p*=0.001). On the other hand, carriage-negative participants’ levels of 15B CPS IgG at day 14 were comparable to those at baseline (median 3681 ng/ml at baseline vs. 3037 ng/ml at day 14, *p*= 0.64) (Figure 4A).

### Baseline levels of 15B CPS-specific memory B cells did not associate with protection against carriage acquisition

We previously found that baseline levels of IgG secreting memory B cells (B_MEM_) against the capsular polysaccharide (CPS) of the inoculated strains correlate with protection against experimental colonisation with 6B pneumococcus (26). To determine whether this also applied to 15B serotype we measured total levels of B_MEM_ to 15B CPS by ELISpot at baseline (5 days prior to inoculation) and 14 days post-challenge (D14). Levels of 15B-CPS IgG-secreting B_MEM_ cells at baseline were similar in carriage-positive and carriage-negative participants (median: 3.55 [IQR: 2.01-5.50] vs 4.6 [IQR: 2.70-6.00], respectively (*p*=0.53) (Figure 4B). Consistent with our previous findings, post-challenge, carriage-negative individuals had a 2.3 log decrease in levels of total 15B B_MEM_ cells (median 3.55 at baseline to 1.55 at day 14) (*p*=0.008), whereas pneumococcal colonisation did not lead to a substantial change of these levels in the carriage-positive group compared to baseline levels (4.6 at baseline to 3.4 at day 14) (*p*=0.55) (Figure 4B).

### Baseline serum levels of anti-protein IgG to 75 pneumococcal antigens did not associate with protection against 15B

No significant association was observed between baseline serum IgG levels to any proteins in a panel of 75 protein antigens tested and protection against 15B colonisation (Figure 5). However, as shown in Figure 5, a trend was seen where carriage negative participants had higher levels of IgG at baseline against numerous proteins.

**Figure 5.**
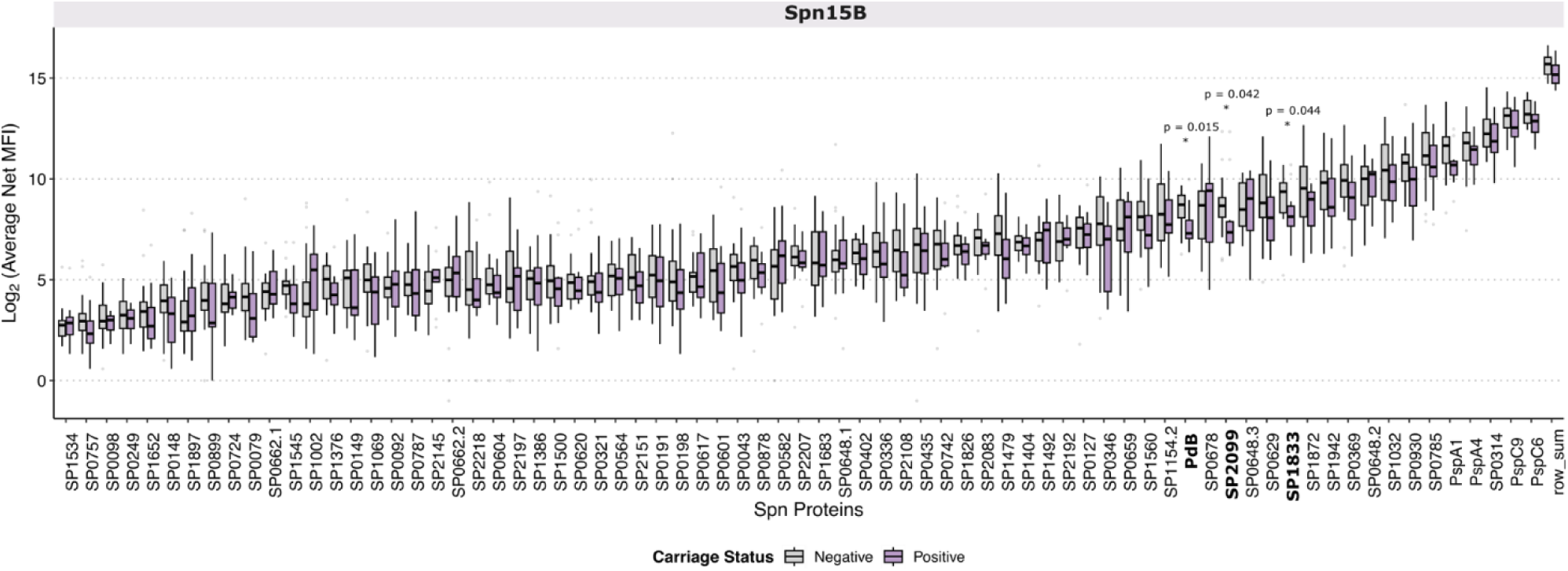
Baseline serum IgG responses to pneumococcal protein antigens prior to 15B challenge. Baseline levels of serum IgG against 75 pneumococcal proteins in study participants, measured as average net median fluorescence intensity (MFI). Bars indicate interquartile range. 15B carriage-negative (grey, n=22), 15B carriage-positive (purple, n=11).

Due to sample size limitations, we did not have the statistical power to determine significant changes in circulating anti-protein IgG response post-challenge with 15B as compared to baseline. However, we observed that fold-change in anti-protein IgG was generally reduced 14 days after challenge in the carriage-negative cohort but increased in the carriage-positive cohort (Figure 5).

Protein-specific IgG levels did not differ significantly five days prior to heterologous re-challenge with 6B between participants who were subsequently colonised by 6B and those who were not (Figure 6). Furthermore, fold change in IgG levels at this time point from those observed 14 days following the initial 15B challenge were comparable across both cohorts (Figure 7).

**Figure 6.**
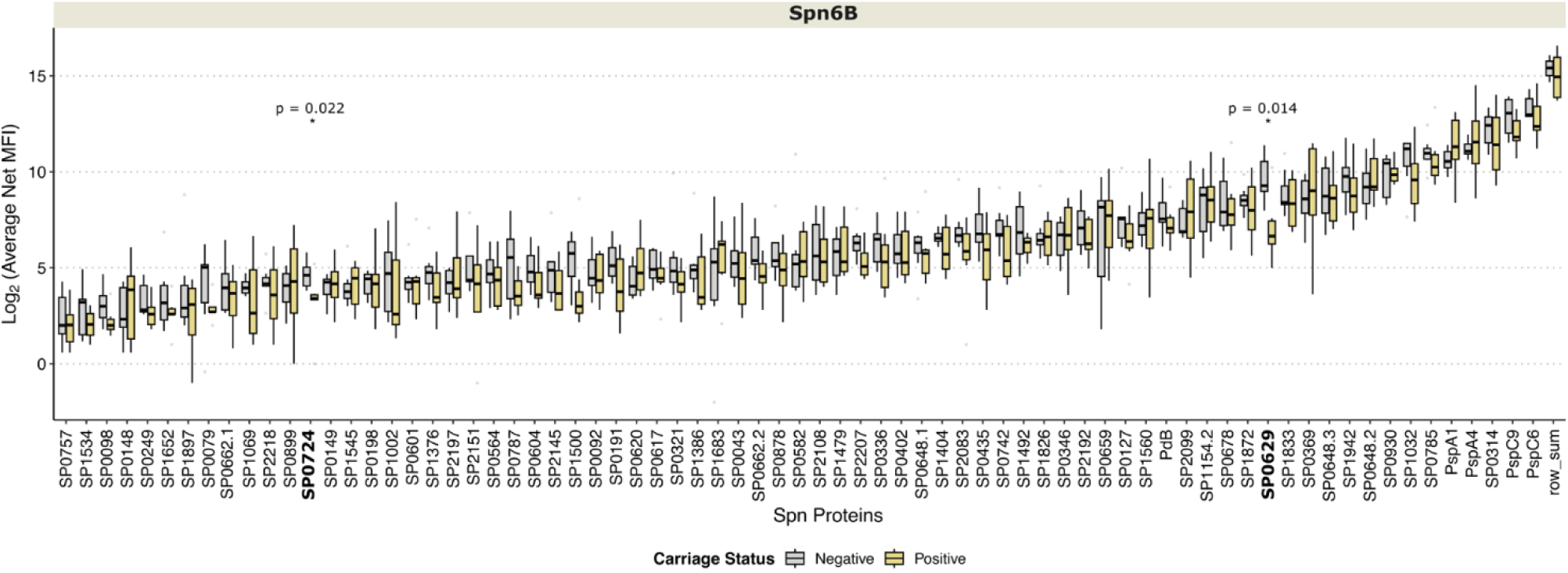
Baseline serum IgG responses to pneumococcal protein antigens prior to 6B rechallenge. Serum IgG levels against 75 pneumococcal proteins were measured in study participants at the pre-rechallenge time point, following prior inoculation and carriage with 15B. Participants were stratified based on 6B carriage status: carriage-negative (grey, n=7) and carriage-positive (yellow, n=7). Responses are shown as average net median fluorescence intensity (MFI), with bars indicating the interquartile range.

**Figure 7.**
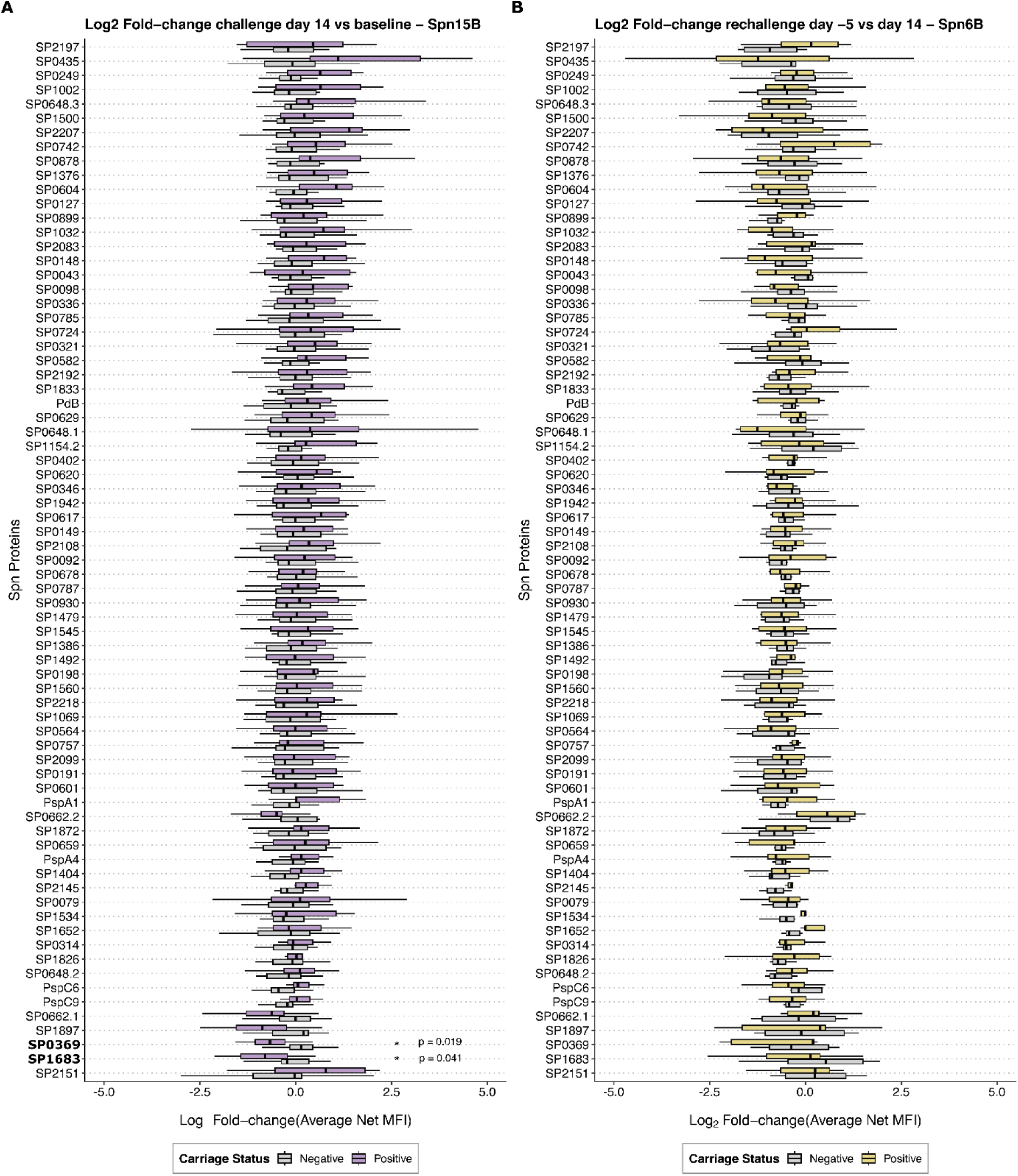
Fold changes in serum anti-protein IgG associated with carriage following experimental pneumococcal challenge (15B) and re-challenge (6B). (A) Fold change in anti-protein serum IgG (measured as average net median fluorescence intensity, MFI) from baseline in carriage-positive (n=9) and carriage-negative volunteers (n=15) following experimental challenge with 15B. Post-challenge samples were taken 14 days after initial challenge. Bars indicate interquartile range. (B) Fold change in anti-protein serum IgG (expressed as mean net median fluorescence intensity [MFI]) measured 5 days prior to challenge with 6B (6–9 months after the initial 15B challenge), relative to levels at 14 days following the initial 15B challenge. Data are shown for participants who developed carriage after re-challenge (positive, n=7) and those who did not (negative, n=7). Bars represent the interquartile range.

### Colonisation with serotype 15B pneumococcus did not associate with protection against acquisition of serotype 6B

Across our CHIM studies, in which adults of the same age range were inoculated with the same dose of serotype 6B, carriage rates consistently ranged from 39-58%, with an average colonisation rate of 47.7% (115/241) (11, 31–34).

Here, we investigated whether a recent (up to 9 months) colonisation episode with the 15B strain could confer heterologous protection against the 6B strain (Supplementary Table 2). No participant was naturally colonised with pneumococcus at the time of the re-challenge. Six out of 13 participants previously colonised with 15B became colonised with 6B following re-challenge (46.2%), consistent with the average colonisation rate previously achieved with 6B in our CHIM studies (11, 31–34).

### Cytokine responses to pneumococcal challenge

To assess whether carriage induced nasal tissue mucosa responses to a heterologous pneumococcal strain, nasal micro-biopsies were collected from a subgroup of volunteers (n=14) at baseline and 14 days post inoculation with 15B. All had been inoculated with 8 x 10^4^ CFU per naris; 7 were carriage-positive and 7 were carriage-negative. Nasal cells were stimulated *in vitro* with heat-killed 15B, heat-killed 6B or RPMI cell culture medium (control). We measured levels of a selected group of 6 cytokines (IL6, IL10, GM-CSF, TNFα, MIP-1β and MIP-1α) in the nasal cell supernatant following stimulation. These cytokines were previously selected for their role in the control of experimentally induced pneumococcal colonisation in humans (27). The fold change (FC) in cytokine concentrations following stimulation with 15B and 6B compared with mock stimulation with RPMI cell culture medium is illustrated in Figure 8.

**Figure 8.**
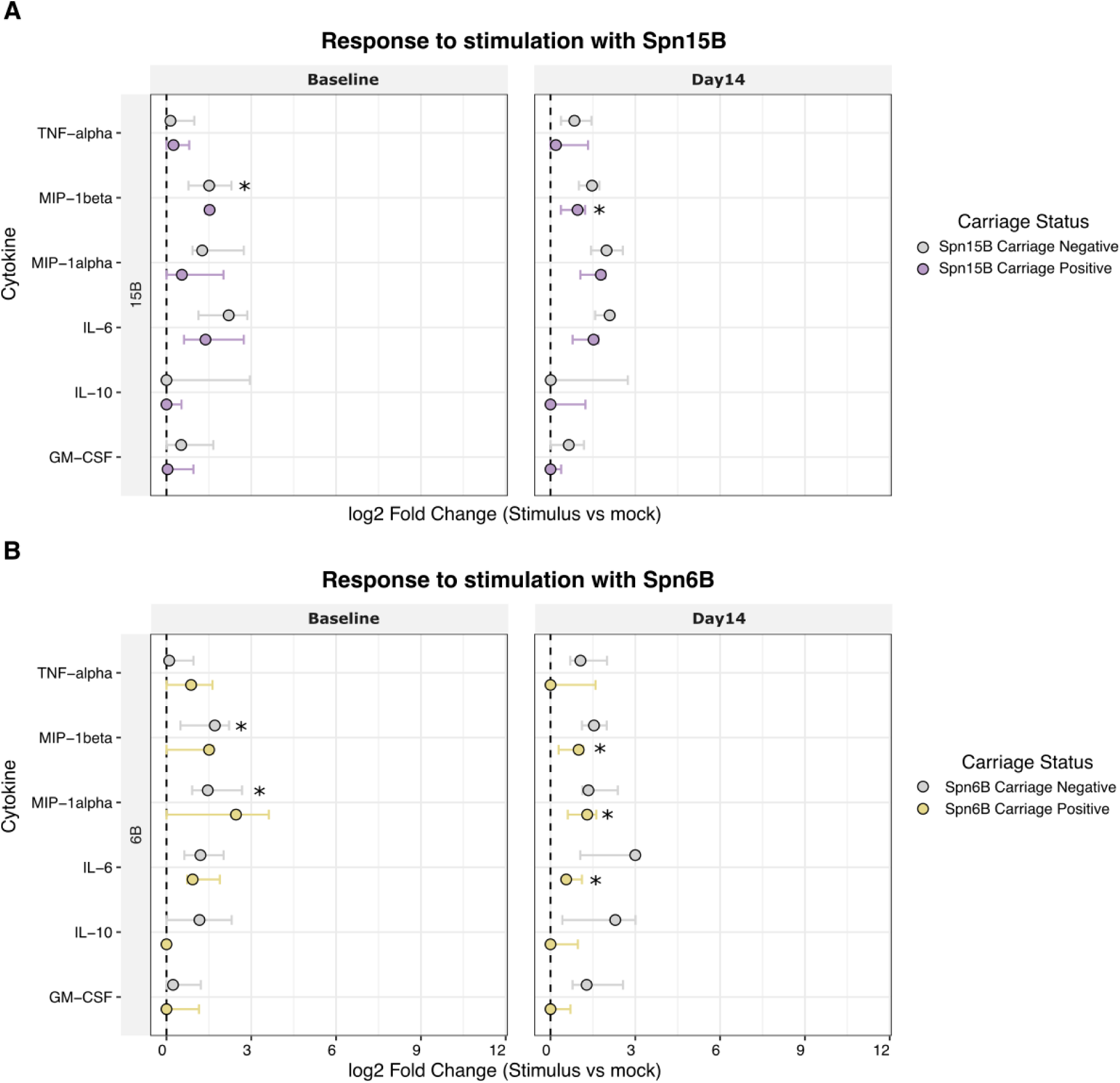
Fold changes in cytokine responses to pneumococcal stimulation in nasal cells stratified by carriage status before and after experimental colonisation. (A) Log₂ fold change in cytokine production following stimulation with heat-killed 15B (Spn15B) compared to mock (RPMI) in carriage-positive and carriage-negative participants. (B) Log₂ fold change in cytokine production following stimulation with heat-killed 6B (Spn6B) compared to mock (RPMI) in carriage-positive and carriage-negative participants. Nasal cells were collected 5 days prior to inoculation (Day −5) and 14 days post-inoculation (Day 14) with 15B. Points represent median values, and error bars indicate the interquartile range (IQR). The dashed vertical line denotes no change relative to mock stimulation (log₂ fold change = 0). Asterisks indicate statistically significant differences between carriage groups (p < 0.05).

At baseline (before intranasal challenge with 15B), a significant change in MIP-1β concentration following *in vitro* stimulation with 15B and 6B compared with mock stimulation was detected in participants who remained negative (n=7) (15B; FC=2.60, *p*=0.03, 6B; FC=3.34, *p*=0.03) but not in those who were susceptible to colonisation. The same was seen for MIP-1 α concentrations but only following *in vitro* stimulation with 6B (n=7) (FC=2.81, *p*=0.03). MIP-1 β (CCL4) and MIP-1*α* (CCL3) are chemoattractants and activators for immune cells, such as monocytes and T cells, CD4^+^ T cells and CD8+ T cells, respectively. There was no significant response observed in any of the other cytokines tested.

In cells collected 14 days post inoculation with 15B, a significant induction in MIP-1β concentration in response to restimulation with 15B was seen in the carriage-positive group (FC= 1.65, *p*= 0.05) but not the carriage-negative group. Furthermore, MIP-1α (FC= 2.00, *p*= 0.03), IL-6 (FC= 1.47, *p*= 0.03) and MIP-1 β (FC= 1.99, *p*= 0.02) levels were all significantly increased in response to stimulation with 6B in the cohort with established 15B colonisation.

## Discussion

We report the development of a new pneumococcal CHIM to investigate the colonisation dynamics of pneumococcal serotype 15B. We have shown that 8 × 10^4^ CFU per naris is the optimum inoculation dose for 15B experimental challenge. In contrast to serotypes 3 and 6B, which have demonstrated the ability to achieve a carriage rate of up to 70 and 60% with an inoculum dose of 8 × 10^4^ CFU per naris (11, 35), attack rates with 15B peaked at 40% at that dose, and increasing inoculum doses for 15B led to poorer colonisation rates (11%). When comparing carriage densities measured by classical microbiology, the density of 15B was also lower than that of 6B and serotype 3 strains (35).

Data generated *in vitro* demonstrated that 15B is more likely to bind to the surface of epithelial cells, while 6B is more likely to be internalised by them. Micro-invasion of the epithelial barrier occurs during colonisation and induces inflammation and innate immune responses (29). We hypothesise that the increased epithelial cell association and reduced microinvasive capacity of 15B relative to 6B results in greater exposure to innate defence mechanisms in the upper airway, thereby limiting its colonisation potential.

Experimental colonisation with 15B did not associate with protection against heterologous re-challenge with 6B, which resulted in a 46.2% colonisation rate, consistent with our observations from previous 6B challenge studies (11, 31–34). Similar to findings of prior 6B studies (11), a significant increase in serum IgG levels to the 15B CPS was observed following colonisation but no relationship was found between baseline IgG levels and colonisation outcome following initial challenge (11, 26). Similar levels of 15B CPS IgG-secreting B_MEM_ in peripheral blood were found at baseline in carriage-positive and carriage-negative volunteers. Previous work from our group has shown that volunteers protected against carriage acquisition of 6B had approximately three-times higher levels of CPS 6B-specific B_MEM_ at baseline compared with those who went on to carry the bacteria (26). Thus, our data may indicate that the level of B_MEM_ required for protection against colonisation differs amongst pneumococcal serotypes. Our findings may also reflect variation in both the timing of colonisation with serotype 15B and the persistence of the resulting immune responses, as memory B-cell maintenance may differ across polysaccharide serotypes. Alternatively, the discrepancy may be due to the use of frozen cells in this work rather than fresh PBMC samples previously used for the 6B work (26). Notably, the levels of circulating CPS 15B B_MEM_ population significantly declined post challenge in the carriage-negative group, suggesting a potential role of CPS 15B-specific B_MEM_ cells in protection against carriage acquisition. We have previously observed that the number of B cells decrease in the nose following exposure to pneumococcus (carriage-negative) (36). The role of nasal tissue capsular-specific resident memory B cells in protection and whether they increase by PCV vaccination needs to be further investigated.

Furthermore, no increase in serum IgG levels to 15B capsular CPS was found following intranasal exposure to 15B serotype that did not result in detectable colonisation. Conversely, in participants who acquired experimental 15B colonisation during the study, a significant increase in serum IgG levels to 15B capsular PS was observed. These findings are in line with our previous studies, in which established colonisation with 6B increased IgG responses to CPS and multiple protein antigens but challenge without colonisation did not associated with a change in CPS 6B specific IgG levels in serum (12, 31). For serotype 3 challenge studies, colonisation was also required to increase IgG levels to CPS 3 (35).

Baseline levels of anti-protein IgG in serum were not predictive of protection against experimental colonisation, consistent with previous findings that anti-protein antibodies at baseline do not correlate with protection against experimental carriage with 6B in young or older adults (11, 32, 37).

No increase in circulating anti-protein IgG was detected at day 14 post-15B inoculation in either carriage-positive or carriage-negative groups. It is unclear whether a more robust anti-protein IgG response may have been detected at a later timepoint in volunteers colonised with 15B. A significant systemic anti-protein IgG response to colonisation has previously been detected at 14 days post initial challenge with 6B (11), suggesting that 15B colonisation may indeed be less immunogenic than 6B. Mucosal anti-PspA IgG response to colonisation with 6B however has been shown to reach significantly higher levels as compared to baseline only at 28 days post-challenge but not at 14 days post-challenge (11). Our low sample size and the sampling at 14 rather than day 28 may therefore have limited our power to detect a significant rise in circulating anti-protein IgG.

Cytokine analysis revealed elevated MIP-1β at baseline in carriage-negative participants and post-challenge in carriage-positive volunteers, suggesting a role for MIP-1β in control of colonisation with 15B. Analogous responses were seen with MIP-1α following *in vitro* stimulation with 6B. Previous work has found a significant increase in production of IL6, IL10, GM CSF, TNFα and MIP-1α following 6B stimulation (27). This study also found that the increase in these cytokines correlated with a reduction in pneumococcal density in the nasopharynx (27), supporting serotype-specific differences in innate immune activation. MIP-1α and MIP-1β are proinflammatory and chemotactic cytokines that recruit macrophages, neutrophils and certain lymphocytes to sites of infection (38).

Epidemiological data indicate both serotype-specific and serotype-independent immune responses develop following pneumococcal colonisation (39, 40). The parallel age-related decline in invasive pneumococcal disease across both common and uncommon serotypes supports an important role for serotype-independent protection. Despite only modest age-related increases in anti-capsular antibody levels, incidence of distinct serotypes closely tracks one another throughout childhood, suggesting the contribution of non-capsular immune mechanisms to natural protection against invasive disease (41). Possible mechanisms include CD4 Th17 memory cells, trained innate immune responses and antibodies against non-serotype specific pneumococcal proteins (25, 42, 43).

A more detailed machine-learning analysis by our group has identified potential serotype-independent correlates of protection against colonisation, pointing out the potential importance of combined antibody and T cells responses to proteins (25).

A significant strength of this study is the use of our unique experimental human pneumococcal challenge and re-challenge model. The model allows for the study of pneumococcal colonisation following a known exposure event to robustly characterise carriage status as well as the duration and density of any carriage episode and compare serotypes as well as correlates of protection.

Our study is limited by the absence of a contemporaneous control group in the re-challenge cohort, precluding direct comparison of 6B colonisation rates between volunteers previously colonised with 15B and those naturally protected from experimental colonisation. However, our extensive data from studies conducted over the past decade in over 1500 participants inoculated with 6B pneumococcus indicate expected 6B colonisation rates of 39–58%, with an average colonisation rate of 47.7% (11, 31–34), which are comparable to the 46.2% colonisation rate observed in the present re-challenge cohort. Future work will address this limitation by concurrently challenging carriage-negative and carriage-positive cohorts to enable direct assessment of heterologous re-challenge outcomes.

In addition, the limited sample size and duration of follow-up may have reduced our ability to detect immune responses that could become apparent in larger cohorts or with longer follow-up. We previously demonstrated that experimental colonisation rates with 6B differ between sexes (44). In the present study, we did not disaggregate data by sex due to limited sample sizes. However, we anticipate that larger datasets will enable investigation of sex as a biological variable, potentially revealing immune differences between sexes.

Our findings emphasise that natural colonisation alone may be insufficient to induce optimal and durable heterologous protective immunity across pneumococcal serotypes. However, the consistent observation across our CHIM studies that approximately 40–50% of healthy adults resist experimental colonisation suggests that pre-existing mucosal immunity may play a critical role in determining susceptibility. Adults are repeatedly exposed to pneumococcus throughout life and are therefore likely to harbour pneumococcal-specific memory T cells, including tissue-resident memory populations within the nasal mucosa. In this context, the quality, magnitude and localisation of the mucosal innate response appear critical. The nasal activation signatures we observed here and in our previous work, particularly MIP-1α, MIP-1β and IL-6, are consistent with recruitment and activation of monocytes and memory T cells. Such chemokine-mediated cellular orchestration may represent an early checkpoint in determining colonisation outcome. Individuals resistant to carriage display differential and heightened baseline responsiveness, suggesting that differences in innate immune set-point and mucosal immune readiness may influence whether bacterial inoculum is rapidly cleared or progresses to stable colonisation.

Recent work by the Pulendran group and colleagues (45) has demonstrated that mucosal vaccination can harness pre-existing antigen-specific memory T cells to reprogramme airway innate cells and generate antigen-agnostic protection against diverse respiratory pathogens. In contrast to the durable innate imprinting described in murine vaccination models, natural pneumococcal colonisation in humans may induce a magnitude or persistence of mucosal immunity that is insufficient for robust heterologous protection. It is plausible that experimental colonisation exposes underlying heterogeneity in mucosal immune readiness rather than simply reflecting serotype-specific immunogenicity. Together, these findings suggest that deliberate mucosal immune priming, designed to amplify activation of pre-existing pneumococcal-specific anti-protein T cells, recruitment of new cells, and enhancement of innate-T cell cross talk, could represent a rational strategy to enhance serotype-independent protection against colonisation. Such approaches may complement current conjugate vaccines by targeting the mucosal niche where transmission begins and by promoting broader strain-transcending immunity.

## Author contributions

Conceived the study – EM, DMF, JRy

Data acquisition – VC, EM, KSC, ELG, CMW

Generated the ELISpot and ELISA data – EM

Generated the Luminex data – KSC

Generated the colonisation data – ELG, EN, SP

Generated the colonisation data adhesion/invasion assay data – CMW

Performed data analysis – VC, EM, KSC, ELG, CMW

Scientific discussion – EM, KSC, ELG, CS, BU, DMF, JRy, RM, SBG

Provided protein library – RM, YJL, ENM, RT

Generated all Figures – PGD, EM, KSC

VC, EM, KSC, ELG jointly interpreted the results and wrote the manuscript with contributions from all co-authors. All authors read and approved the final manuscript.

## Acknowledgements

The authors thank the respiratory research team members: Catherine Lowe, Catherine Molloy, Kelly Convey, Rachael Robinson and Stephanie Griffiths. Trial Steering Committee members: Professor Neil French, and clinicians who provided safety on-call cover Dr Ben Morton, Dr John Blakey, Dr Steve Aston and Prof. Neil French and the Liverpool School of Tropical Medicine (LSTM) Respiratory group. We thank the late Dr Hassan Burhan for his clinical support. We also thank Dr D. Cleary, University of Southampton for the donation of the 15B serotype used in this study. The study was co-sponsored by the Royal Liverpool and Broadgreen University Hospital Trust (RLBUHT) and Liverpool School of Tropical Medicine (LSTM).

