## Supplementary Figure 1 for "Innate immunity associates with protection from pneumococcal colonisation, but colonisation does not confer capsule-independent protection"

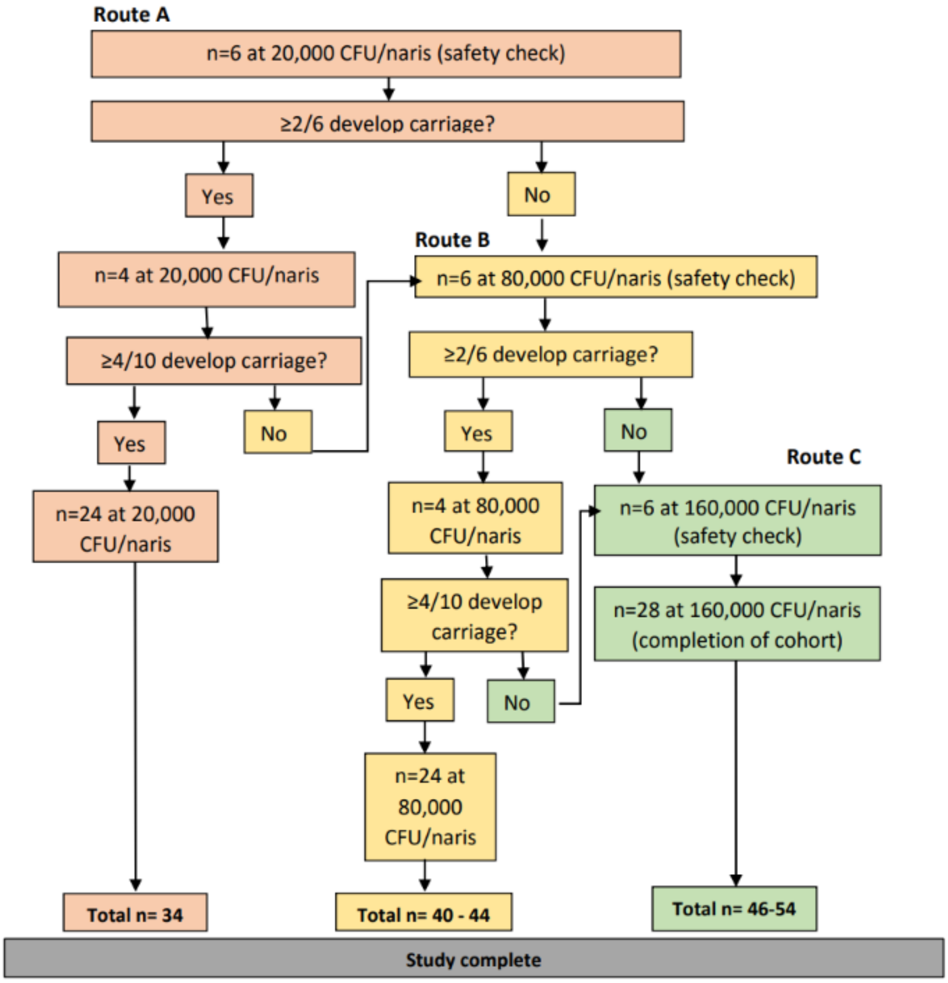


***Supplementary Figure 1: Dose ranging and reproducibility study design, adapted from (1).*** We initially challenged 10 participants with 20,000 CFU/naris based on carriage rates found previously for a serotype 23F pneumococcal isolate (2). This dose was also important to check safety of strain inoculation. These individuals were monitored for adverse effects and for pneumococcal carriage. If the carriage rate had been ≥4/10 (Dose ranging cohort A), a further cohort of 24 participants would have been challenged to complete a total of 34 participants (Reproducibility cohort). As carriage rate was insufficient, the algorithm was restarted with a higher challenge dose (Dose ranging cohort B with 80,000 CFU/naris then dose ranging cohort C with 160,000CFU/naris).
