## Supplementary Table 2 for "Innate immunity associates with protection from pneumococcal colonisation, but colonisation does not confer capsule-independent protection"

***Supplementary Table 2:*** Participants of re-challenge cohort (6B challenge)

| Cohort | Number of subjects | Age (yr) | Sex (M:F) | Study period | Dose (CFU/naris) |
| --- | --- | --- | --- | --- | --- |
| **Re-challenge** | 13 | 31.38 ± 12.18 | 4:9 | Feb-Jul 2018 | $8.5 \times 10^4 \pm 4343$ |

*Values are means ±SD*
