## Supplementary Figure 2 for "Innate immunity associates with protection from pneumococcal colonisation, but colonisation does not confer capsule-independent protection"

A) B)

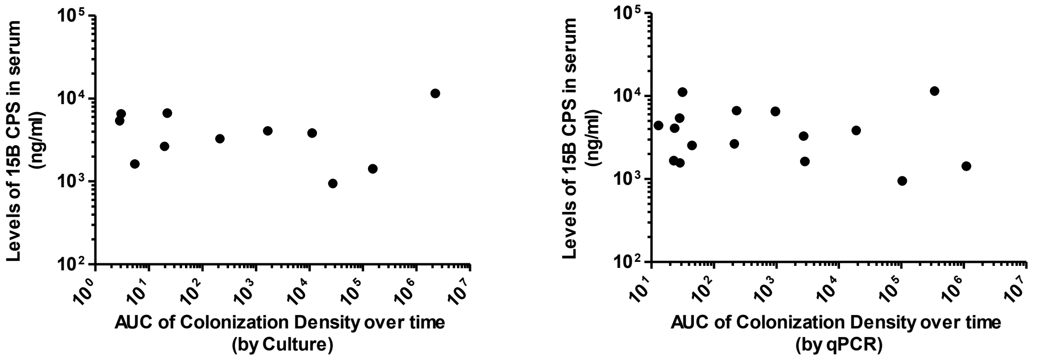


***Supplementary Figure 2:*** Correlation between baseline 15B CPS IgG levels and area under the curve of colonisation density over time detected by A- classical microbiology and B- qPCR. Culture; $R_S$= -0.21 spearman's rank correlation coefficient, P=0.54. qPCR; $R_S$=-0.082 spearman's rank correlation coefficient, P=0.76.
