## Supplementary Table 1 for "Innate immunity associates with protection from pneumococcal colonisation, but colonisation does not confer capsule-independent protection"

***Supplementary Table 1:*** Participant demographics for dose ranging and extended cohorts inoculated with serotype 15B pneumococcus.

| Cohort | Number of subjects | Age (yr) | Sex (M:F) | Study period | Dose (CFU/naris) |
|---|---|---|---|---|---|
| **Dose ranging A** | 10 | 29 ± 10.25 | 4:6 | Sep 2017 | 1.9 x$10^4$ ± 997 |
| **Dose ranging B** | 10 | 31 ±9.76 | 3:7 | Oct 2017 | 7.3 x$10^4$ ± 1581 |
| **Dose ranging C** | 9 | 21 ± 3.03 | 4:5 | Nov2017 | 1.9 x$10^5$ ± 0 |
| **Reproducibility** | 25 | 21.68 ±4.48 | 12:13 | Nov-Dec2017 | 9.5 x$10^4$ ± 10696 |

*Values are means ±SD*
